# Men and sexual and reproductive healthcare in the Nordic countries: a scoping review

**DOI:** 10.1101/2021.04.20.21255777

**Authors:** Mazen Baroudi, Jon Petter Stoor, Hanna Blåhed, Kerstin Edin, Anna-Karin Hurtig

## Abstract

**Context:** Men generally seek healthcare less often than women and, other than traditional gender norms, less is known about the explanation. The aim was to identify knowledge gaps and factors influencing men regarding sexual and reproductive healthcare (SRHC) in the Nordic countries.

**Methods:** We searched PubMed and SveMed+ for peer-reviewed articles published between 2010 and 2020. The analyses identified factors influencing men’s experiences of and access to SRHC.

**Results:** The majority of the 68 articles included focused on pregnancy, birth, infertility, and sexually transmitted infections including HIV. During pregnancy and childbirth, men were treated as accompanying partners rather than individuals with their own needs. The knowledge and attitudes of healthcare providers were crucial for their ability to provide SRHC and for the experiences of men. Organizational obstacles, such as women-centred SRHC and no assigned profession, hindered men’s access to SRHC. Lastly, the literature rarely discussed the impact of health policies on men’s access to SRHC.

**Conclusions:** The identified knowledge gap indicates the necessity of the improved health and medical education of healthcare providers, as well as of health system interventions.

## Introduction

Men generally seek healthcare, especially primary healthcare, to a lower degree than women, and this also applies to sexual and reproductive healthcare (SRHC).^1,2^ For example, 23% of over 40 year-old men in Europe reported sexual dysfunction but only one-quarter of them sought healthcare,^3^ and similar results have been reported in Sweden.^4^ Also, studies from Sweden and Norway have indicated that youth clinics are perceived as “women clinics”. Therefore, fewer men seek these services compared to young women.^5,6^ Additionally, men test themselves for sexually transmitted diseases to a lower extent compared to women.^7,8^ However, various groups of men might have different health seeking behaviours. For example, men with high socioeconomic status^1,9^ and men who have sex with men (MSM) seek SRHC more often.^10,11^

Traditional gender norms might urge men to be independent, strong and invulnerable and also hinder them from acknowledging having problems, creating a barrier to seeking healthcare.^12^ In particular, admitting sexual health problems might imply more vulnerability for men, thus decreasing the likelihood of seeking healthcare.^5,13^ Even though gender norms play an important role in men’s health seeking behaviours, it cannot alone explain the lower utilization of SRHC. Men who eventually sought SRHC did not get the help they expected. For example, more than half of men who sought help related to sexual function in Sweden reported not getting enough support.^4^ Furthermore, men often felt excluded in healthcare related to infertility and pregnancy.^14,15^ This mirrors the lack of response of the health system to men’s needs that can be related to healthcare organization and delivery,^9^ including no support or guidelines for health professionals to promote men’s SRH.^16^ Additionally, health and medical education in Sweden, as an example, does not have enough focus on men’s SRH.^16,17^

The Nordic countries are among the best in the world in the available international gender equality statistics.^18^ Since gender inequality affects women’s sexual and reproductive health (SRH) to a larger degree compared to men,^19,20^ there is a greater focus on women’s rights to SRH. Men’s SRH does not get the same attention in practice and little is known about men’s SRH in the Nordic countries.^21^ The available literature mainly focuses on gender norms and masculinities and its link to health seeking behaviours and risk taking, while much less is known on how men are experiencing SRHC.^9,21-23^

### Aim

The aim of this scoping review was to identify knowledge gaps and factors influencing men regarding SRHC in the Nordic countries during the period between 2010 and 2020.

## Method

### Search strategies and selection criteria

A structured search of the literature was conducted using two databases, PubMed and SveMed+ (a Scandinavian database). Search terms included sexual and reproductive health, men, healthcare, experiences, and Nordic countries (see Appendix 1 for detailed search terms). The following eligibility criteria were used: (1) peer-reviewed empirical studies, all study designs were considered; (2) published between January 2010 and May 2020; (3) assessing men’s experiences in SRHC or perspectives of HCPs on men’s SRHC; and (4) conducted in the Nordic countries.

The initial search gave 1286 articles (896 from PubMed and 390 from SveMed+). After screening the titles and abstracts, 108 articles were read in full, and after being judged for their eligibility, 44 articles remained. An additional 24 papers were identified through the reference lists of these papers, resulting in 68 papers included in this scoping review (Figure 1). The articles were judged for eligibility by the first author, but when uncertainties arose, two co-authors read and judged the articles for eligibility separately. The three researchers then discussed the articles and decided unanimously on the inclusion/exclusion of these articles.

**Figure 1:**
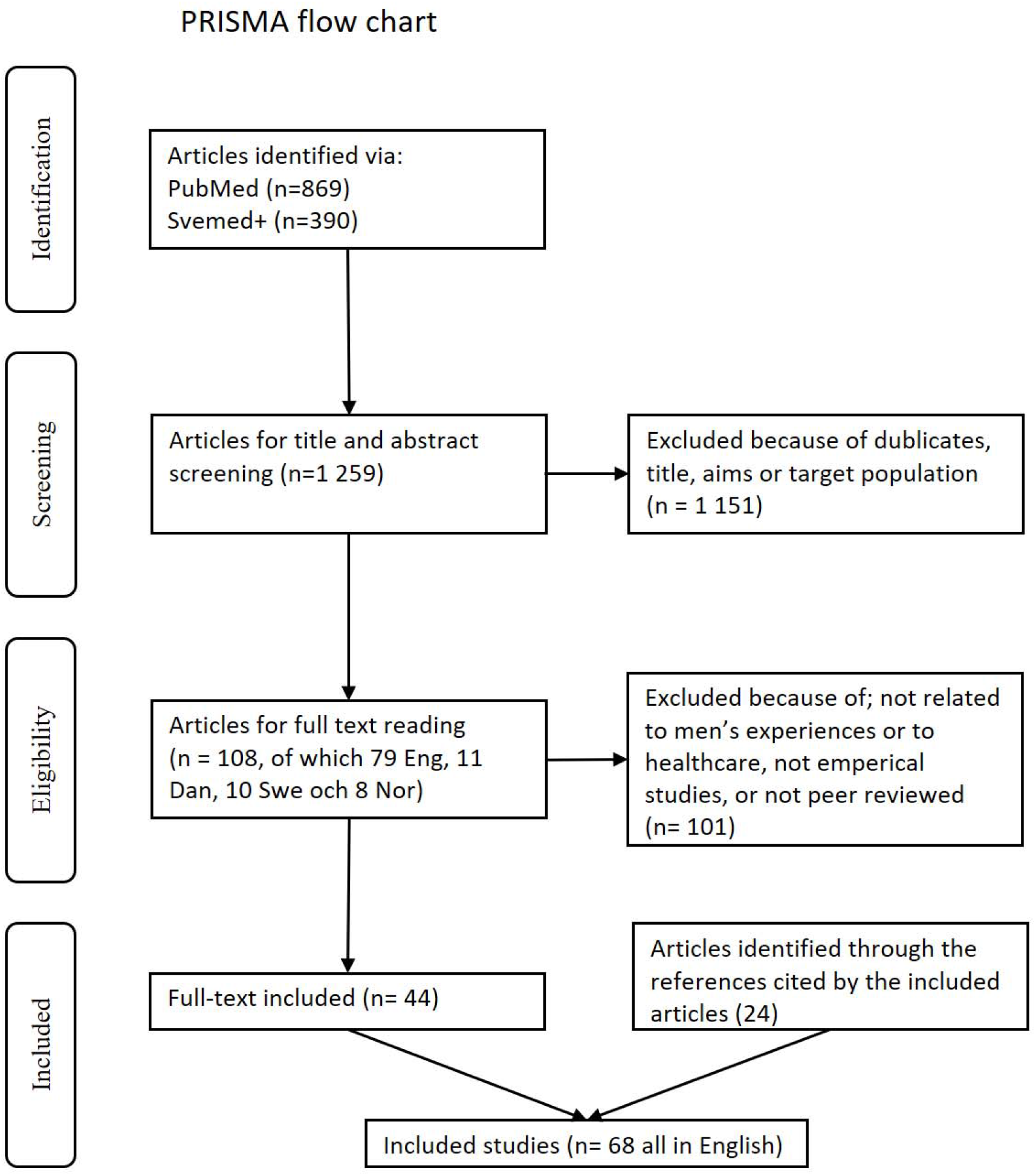
PRISMA flow chart on search results of men’s experiences in sexual and reproductive healthcare in the Nordic countries.

### Data extraction and synthesis

The identified articles were mapped using the World Health Organization (WHO) framework for operationalizing SRH,^24^ and the result parts of each article were extracted and coded using sensitizing concepts of healthcare experiences (Appendix 2). Thereafter, the results were synthesized using a theoretical framework, adapted from Kilbourne et al., which provides health service research perspectives on understanding health and healthcare disparities.^25^

## Results

### Description of the identified studies

Despite not restricting the language of the studies, all the 68 studies included were in English. The absolute majority of the studies were conducted in Sweden (54 articles), while six studies were conducted in Denmark, five in Norway and three in more than one country. No studies were identified from Iceland or Greenland.

Half of the studies (34) adopted a qualitative design, 32 studies a quantitative design and two studies a mixed methods design. Most of the studies (61 articles) were about men’s perspectives of SRHC, while only seven studies covered the perspectives of healthcare providers (HCPs). Of the studies dealing with men’s perspectives, 16 studies assessed women’s perspectives together with that of men. Apart from two articles about the experiences of transgender men, the articles did not mention gender identities. Most of the papers dealing with men’s perspectives referred to the overall experience of healthcare and healthcare staff in general. Of the 28 papers referring to specific primary healthcare providers, 14 mentioned midwives, eight mentioned physicians and six mentioned nurses.

SRH topics were grouped with help of the WHO framework for operationalizing sexual health and its linkages to reproductive health.^24^ The framework demonstrates the interlinked nature between sexual health and reproductive health, yet clearly distinguish topics for intervention and research in both sexual health and reproductive health (Figure 2). Besides the eight topics from this framework, SRH cancers were also added, while the remaining studies with no one topic of focus were grouped under “other”.

**Figure 2:**
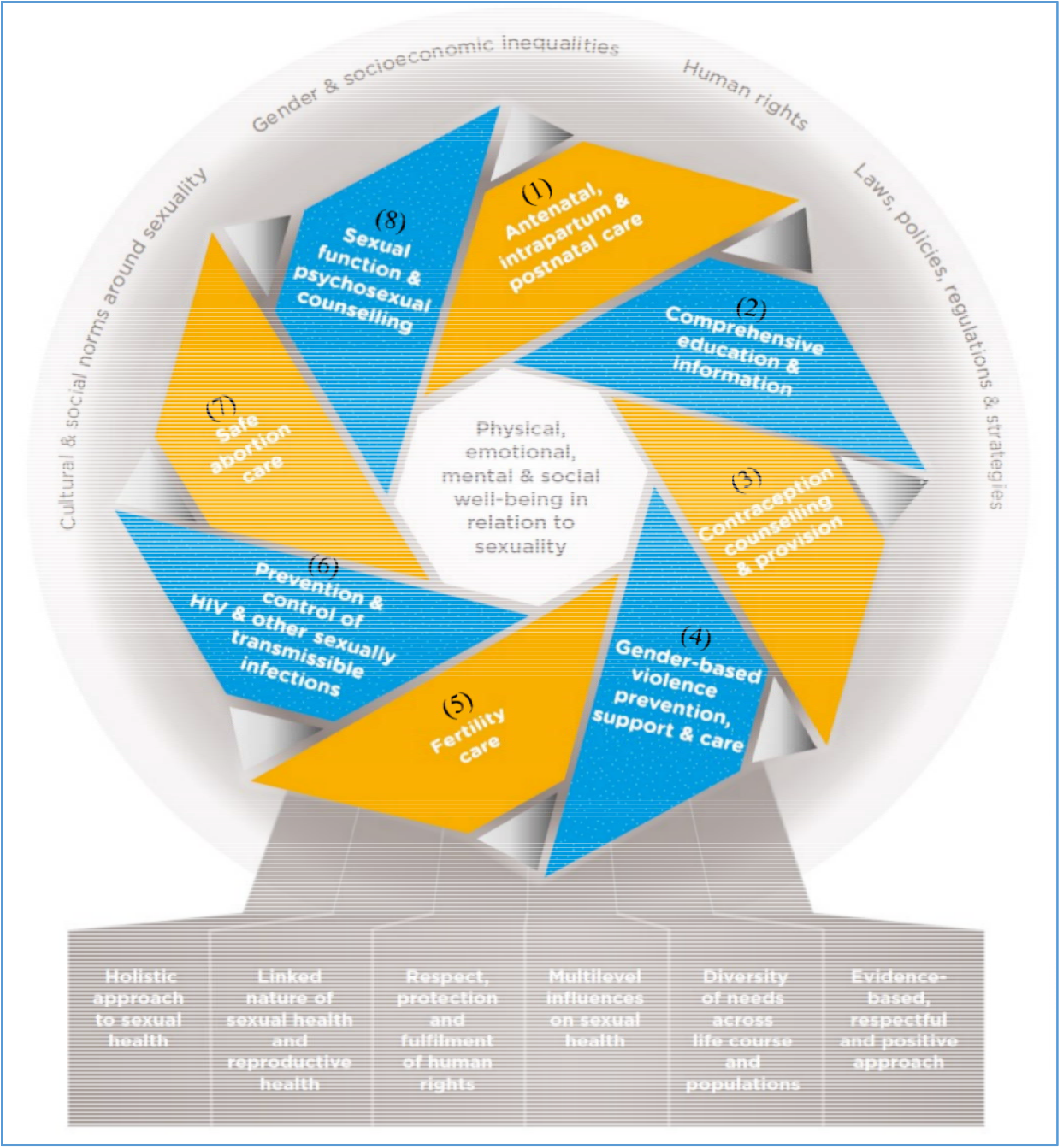
Framework for operationalizing sexual health and its linkages to reproductive health (from “Sexual health and its linkages to reproductive health: an operational approach”).^24^ The intertwined blue and orange ribbons represent sexual health and reproductive health, respectively.

More than one-third of the papers were about the experiences of fathers/expectant fathers during antenatal, intrapartum and postnatal care (25 papers, including 12 about antenatal care and 11 about intrapartum care), while 15 papers dealt with sexually transmitted infections (STIs), mainly HIV (12 papers) and MSM (nine papers). We found 11 papers concerning men’s experiences in infertility care (three of them were related to infertility among cancer patients) and eleven papers in cancer care. We also found four studies dealing with sexual education and information (two of them related to cancer and the other two related to antenatal care), three studies about abortion care, two studies about sexual violence and two studies about sexual functioning and counselling (both related to cancer patients). We found no study dealing with the provision of men’s contraceptive counselling (Figure 3).

**Figure 3:**
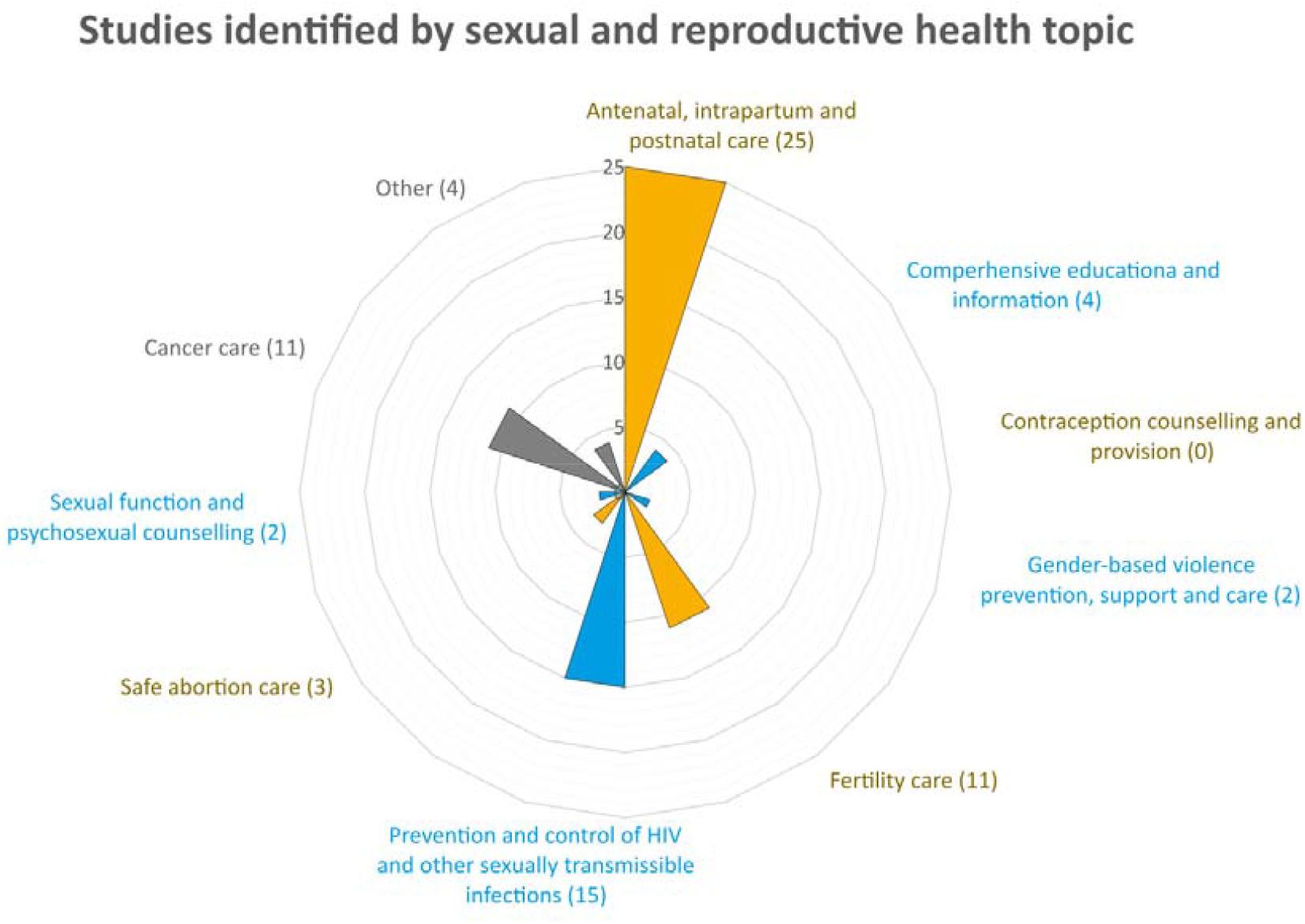
Men’s experiences in sexual and reproductive healthcare in the Nordic countries. Number of studies identified grouped by sexual and reproductive health topics.

### Theoretical framework for analysis

The identified literature dealt with men’s experiences in SRHC from various perspectives and can be organized in the framework adapted from Kilbourne et al.^25^ The factors influencing men’s experiences are divided into (i) individual, including healthcare providers and users; (ii) interpersonal, which deals with the healthcare encounter and contact circumstances; (iii) organizational, which deals with healthcare system factors; and (iv) the larger influence of the community and public policies (Figure 4).

**Figure 4:**
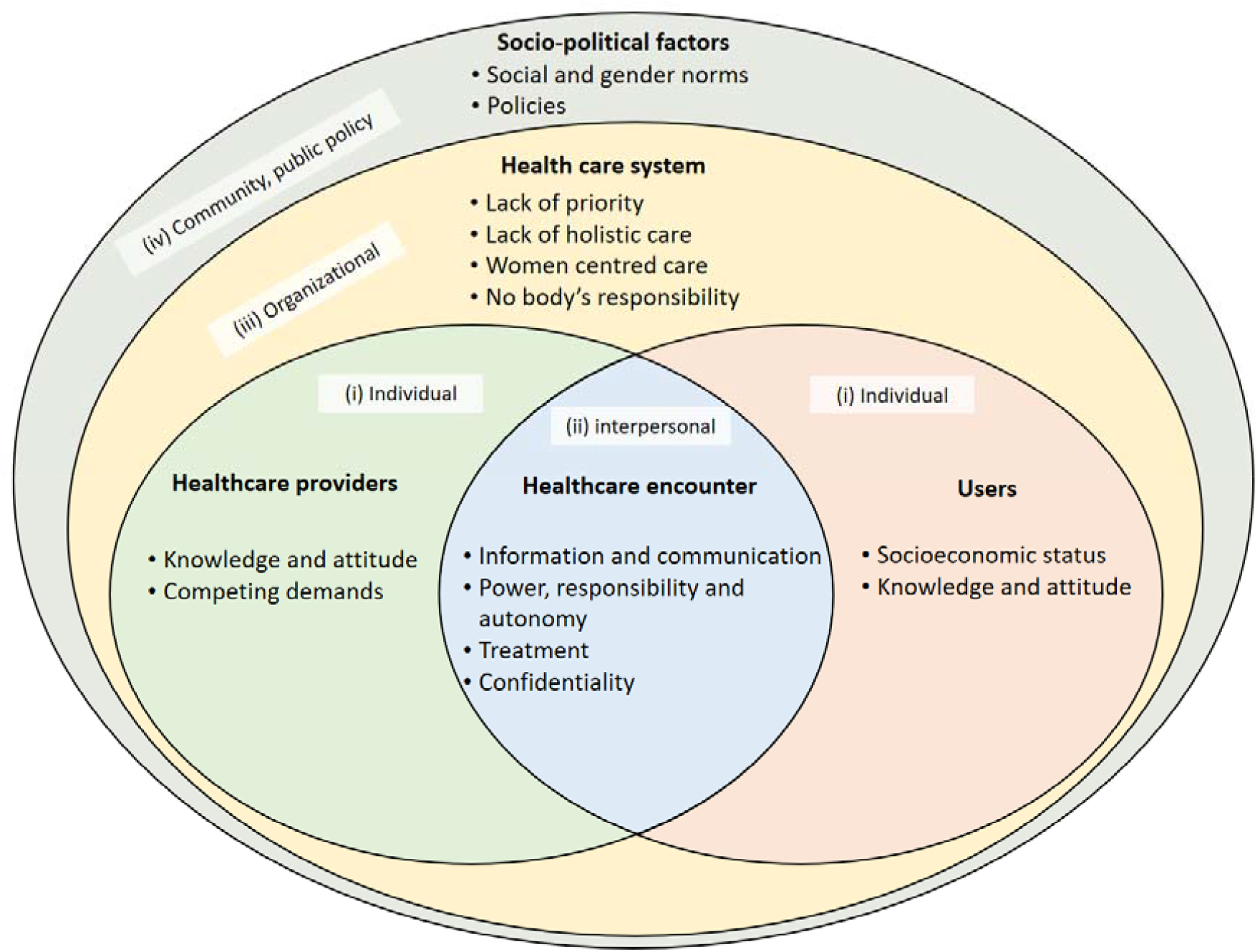
Theoretical framework for analysis of men’s experiences in sexual and reproductive healthcare, adapted from Kilbourne et al.^25^

#### 1. Healthcare providers’ factors

The literature described on how factors related to HCPs, such as sex, attitude, knowledge and competence, affect the HCP-user relationship and experiences of men in healthcare. For example, female HCPs did not prevent men from talking about their concerns regarding infertility.^26^ Similarly, men diagnosed with prostate cancer wished to talk about sexuality with a mature knowledgeable HCP, without considering their sex.^27^ Furthermore, disclosing victimization to female HCPs as compared to male HCPs was claimed to be easier for some men.^28^

##### 1.1. Varied levels of knowledge, competing demands and differing attitudes

Lacking knowledge about men’s SRH was expressed by various HCP professions and were associated with less ability to deal with men’s SRH consultations. For example, nurses perceived that their lack of knowledge was influencing their preparedness to provide sexual health consultations for men.^16^ Midwives also expressed their limited knowledge about male SRH, which was considered essential if inviting men for SRH consultations.^17^ Additionally, less experienced physicians (young and/or under training) felt uncomfortable dealing with sexual health consultations.^29^ Additionally, the competing demands in the form of high workload and limited time hindered HCPs from discussing SRH with men.^16,30,31^

Differing attitudes towards health seeking behaviours of men were found. While most HCPs were described as having positive attitudes, being friendly, sensitive and supportive,^32-35^ some were still perceived as harsh and nonresponsive.^32,33^ These negative attitudes were sometimes perceived by men as discrimination based on their sex, which hindered them, for example, from disclosing victimization.^28^

##### 1.2. The view of men in reproductive healthcare services, an accompanying partner or an individual?

Even though HCPs in reproductive healthcare services usually deal with couples having a common reason for visiting healthcare, in most cases, they have primarily communicated with the women.^26,32,36,37^ Women were in focus during infertility treatment, pregnancy and birth, leading men to feel neglected, invisible and superfluous during the visits.^14,15,38,39^ The lack of interest in listening to or interacting with men also hindered their involvement in supporting their partners, for example, when giving birth.^40,41^

The lack of focus on men might be explained by time constraints and no time being allocated to men’s concerns during visits.^17^ Anyhow, the attitudes and behaviours of HCPs generally made a difference in men’s perception of their involvement or lack of involvement in healthcare.^38^ Couples highlighted the need to treat partners on equal terms and to focus on them as a unit rather than solely on the women,^14,15,26,36^ and they expected communication as inclusive with both partners.^35^ Men also expressed that HCPs should welcome them to more active involvement during birth and support their role as expectant fathers. HCPs should acknowledge men’s needs and give them the opportunity to talk about their concerns.^14,26,32,42^

Examples of good practices involving men in reproductive healthcare are also mentioned in the literature. One example was participatory parental classes or separate parental classes for men and women dealing with men’s concerns related to pregnancy and birth, which helped men to take part and to feel involved.^39,43^ Another example was assigning tasks and continuously informing men during labour and allowing the father to stay at the hospital after the baby is born. These practices gave men a feeling of being important and recognized, hence receiving needed support.^41,44-46^

#### 2. Healthcare users’ factors

The literature described users’ factors that influence their experiences in healthcare. This included men’s socioeconomic situation, including education, age, knowledge and attitude. For example, the ages of users were discussed in relation to the ages of HCPs; nurses were more comfortable talking about sexuality with younger men as compared to men of their own age or older.^16^ Young men, in comparison to young women, were pointed out as being less acquainted with youth clinics or where else to seek SRHC.^5^

Healthcare users’ factors are discussed in the literature mainly in three SRH subject areas, namely, prevention and control of HIV and other STIs, antenatal/ intrapartum care and cancer care, which is elaborated on below.

##### 2.1. Prevention and control of HIV and other STIs

Most of the literature focused on HIV testing and treatment and the sociodemographic factors of users related to it. See Box 1 for more details about the factors discussed in the literature.

###### Box 1: The sociodemographic factors of users in relation to HIV testing and treatment

- **Age:** Younger age was reported to be associated with higher HIV testing among men who have sex with men (MSM)^49,50^ and earlier diagnosis in the general population.^10^
- **Education:** A lower level of education was associated with less testing for HIV in the general population,^50^ but not among MSM.^49^
- **Country of birth:** Studies showed that country of birth was not associated with lower HIV testing among MSM.^49,51^ However, two-thirds of the foreign-born HIV patients had not been tested for HIV at migration to Sweden.^10,52^ Therefore, foreign-born men were more likely to be diagnosed late (65% of foreign-born compared to 43% of Swedish-born) and less likely to optimally adhere to HIV treatment.^10,53^
- **Sexuality:** Since HIV testing was perceived as implicitly implying same sex sexual relations, non-disclosing MSM were more likely to have been never tested for HIV.^50,54^ However, MSM were less likely to be diagnosed late (40% of MSM compared to 67% of heterosexual patients) and less likely to optimally adhere to HIV treatment.^10,53^
- **Knowledge:** Men’s knowledge about HIV transmission was associated with never being tested for HIV among MSM and the general population.^49,50^ Never being tested for HIV was also associated with not knowing if the tests were free or affordable^50,55^ and lack of knowledge about HIV testing services.^44,49,51^ For example, only one-fourth of MSM knew about home sampling (Internet ordered tests),^56^ and around 40% have never heard of the Testpoints programme (peer-led testing performed in MSM clubs, among other places).^57^
- **Risk perception:** The perception of having a very low risk of contracting STIs, including HIV, was highly associated with never being tested for HIV or STIs.^3,7,48,51^

The literature relating to other STIs (besides HIV) was limited to the attitudes of school boys toward HPV vaccination and attitudes of men toward STI testing during the pregnancies of their partners. School boys had a positive attitude with regard to participation in HPV vaccinations; they stated that vaccinating only girls is unfair. Even though they had a positive attitude to share the responsibility of STI prevention, boys rarely used condoms, especially if they knew their sex partner in advance.^47^ Men’s attitudes toward STI testing during pregnancy were diverse. Some men perceived the test as an “infidelity check” that is sensitive and can risk the relationship, while others perceived it as a safety measure that should be “routine” during pregnancy.^13,48^

##### 2.2. Antenatal and intrapartum care

The literature discussed men’s socioeconomic characteristics, knowledge and experiences in antenatal and intrapartum healthcare. Lack of knowledge about antenatal services, such as antenatal classes, was common among men; they usually had not heard about the service before but received information from their partners.^36^ Men who had no social support from family and friends during the pregnancies of their partners were more dissatisfied with antenatal care and less likely to attend parental classes.^58^ Studies found younger age and higher education level were associated with lower satisfaction with the overall birth experience,^40,59^ while no such association was reported in relation to men’s country of birth.^60^ Additionally, younger men as compared to older men, perceived midwives as less supportive, less attentive and as not inspiring confidence.^59^ These differences might be explained by younger men having higher expectations.^61^

##### 2.3. Cancer care and SRH

The literature explored the factors of users related to cancer care, especially the effects of cancer treatment on fertility and sexuality. The majority of physicians claimed that they discussed the impact of cancer treatment on fertility if the patient was at reproductive age. However, one-third of the physicians did not do this regularily.^30,62^ Around half of men in the 41–60 years old age group claimed that they had not received enough information about the effects of cancer treatment on sexual desire, sexual function and fertility.^63^

Similar to other SRH services, lack of knowledge about the services was common, with only around one-fifth of men knowing about the PSA test for prostate cancer screening before testing.^64^ Studies showed no associations between age and the overall satisfaction with cancer care, while a higher level of education was associated with lower overall satisfaction with prostate cancer care.^65^ Furthermore, the literature indicated that manual workers were less likely to receive a bone scan and radical prostatectomy, and they had higher overall and cancer-specific mortalities as compared to non-manual employees.^66^

#### 3. Healthcare encounter factors

The factors under which the healthcare encounters took place influenced the HCP-user relationship and experiences of users. The literature discussed, among other issues, HCP-user communication and the power and autonomy of men.

##### 3.1. Information and communication

Information and communication were recurring themes in all SRH subject areas. More than half of the studies touched upon some aspect of information, or the way it is delivered and communicated. Receiving information was described as valuable and important and made men feel pleased, satisfied and empowered.^52,67-72^ During the birth process, for example, information helped men to feel included and to find their place in supporting their partners and facilitated the decision-making of couples.^32,42,45,67^ Contrarily, lack of sufficient information was associated with more concerns and feelings of exclusion and dissatisfaction.^40,73^ Insufficient information was reported in various healthcare settings, for example, the effects of antenatal care,^36,74-76^ infertility care^69^ and cancer treatment/surgery on sexual health.^63,77^

The literature also discussed the format of information. Oral information was especially preferred when the matter aroused many questions, such as communicating an infertility diagnosis^35^ or HPV vaccination,^47^ while written information was considered more suitable in other cases, such as HIV and STI information for MSM.^70^ However, even though recommended by the National Board of Health and Welfare in Sweden, studies have shown that the majority of men did not receive written information about prostate cancer screening and some were not even aware that they underwent the screening.^64^ In other cases, a combination of oral and written information was considered easier to comprehend, for example, when communicating the side effects of cancer treatment on fertility^34^ (see Box 2).

###### Box 2: The charactersitics of satisfying information and communication - men’s views

- **Clear and simple language:** Clear and proper level information were perceived as important. The inability to understand the medical language of HCPs caused distress. ^32,34,44,67^
- **Reliable:** Contradictions, exaggerations and lack of reliable information caused frustration.^33,42^ Exaggerated information was associated with unease, confusion and a sense of not being taken seriously.^52^ Men wanted to feel welcome when asking questions and wanted honest, consistent and clear answers.^41,42^ Men expressed a need for help to choose reliable websites and organize and discuss the information received.^14^
- **Personalized and relevant:** While general information could be obtained from the Internet, receiving personalized and relevant information from the HCPs was a high priority^26,46,67^ For example, an online patient-nurse communication service played a central role in providing personalized information for cancer patients.^78^
- **Comprehensive and sufficient:** Receiving adequate and comprehensive information was regarded as important.^52^ For example, men highlighted the need for a deeper dialogue about personal experiences or the psychological concequences of male infertility,^26^ as well as psychological support during waiting times for cancer treatment.^77^
- **Appropriate and interactive:** The way HCPs communicated the information affected men’s feelings; a possitive attitude and “a good mood” among HCPs mirrored less stress in men.^73^ Having time to ask questions and interact with HCPs was also appreciated.^31^
- **Timely:** Constant updates of information during their partner’s labour and birth was highly appreciated by men. Men who received timely information felt well informed, calm, secured and satisfied.^32,41,60,73^ On the other hand, receiving information at inappropriate times was perceived as insufficient.^77^
- **Inclusive:** Involving men in the communication as an equal partner in reproductive healthcare was perceived as necessary.^26^

##### 3.2. Lack of control and compromised autonomy in reproductive healthcare

Men’s engagement in reproductive healthcare seemed to be a complex matter; midwives valued men’s involvement, to a certain point, since they experienced over involvement as a possible sign of controlling behaviour or intimate partner violence.^17^ The literature discussed men’s involvement and their lack of control and compromised autonomy in various situations in reproductive healthcare, especially during pregnancy and birth. For example, the inability to help or act during their partner’s birth made men experience lack of power and control.^45,73,79^ Similarly, the uncontrollable process of non-progressing delivery left men with a feeling of helplessness and insecurity.^45^ Men appreciated being involved in the decision regarding their partner’s elective or emergency caesarean section, but 40% of the men felt they were not involved enough.^76,80^ Also, men reported being more in control and more involved in decision-making during an elective caesarean section or normal spontaneous vaginal birth as compared to emergency caesarean section or assisted vaginal birth.^73,76,80^ However, they also described situations where they were forced to participate in tasks and rituals without their consent, (i.e., cutting the umbilical cord or touching the child’s head before the baby was born).^44^ Even though involvement in decision-making during birth was associated with higher satisfaction,^40,76^ it was still important to be able to choose whether to participate or not in different stages of birth.^41^

Compromised autonomy was also reported in the infertility clinic^81^ and when banking sperm before cancer treatment.^31^ To the contrary, control and involvement in decisions were more satisfactory during home abortions. The pregnant woman made the decision, but the partner’s opinion was important for her.^52,71^

##### 3.3. Good treatment increases security and satisfaction

Men wanted HCPs to treat them as persons, respecting their needs, feelings and experiences. HCPs should try to understand the unique situation of each man and take it seriously.^35,41,46,52^ Respectful treatment was highly expected and associated with higher satisfaction with care.^61,82^ It was especially important to deliver negative news with sensitivity.^42,67^ Men who experienced HCPs as professional, empathetic and attentive, felt satisfied, important and “not just a number”.^5,35,50,52^ In other cases, men perceived insensitivity and lack of respect or attention in the comments of HCPs, resulting in feeling disappointed and dissatisfied.^40,52,67^

The support of midwives during antenatal, intrapartum and postnatal care was necessary and created a feeling of security and satisfaction. Providing attention and information and addressing men’s needs and questions helped men to build trust in the midwives and be supportive to their partners.^41,46,60,73-76^ However, men were not always satisfied with the support of midwives, which made men feel insecure, helpless and worried.^40,76,79^

##### 3.4. Confidentiality, a prerequisite to access to SRHC

Confidentiality was considered an essential condition to access certain SRH services, including youth clinics and HIV testing. For example, fear of being recognized in the clinic was one of the main reasons for not being tested for HIV.^49^ Getting an HIV test was considered as implicitly disclosing same-sex sexuality, which led to preferring self-testing as an anonymous alternative, especially among non-gay MSM and those who had never been tested for HIV.^54^ Therefore, anonymous HIV testing outside the healthcare system were requested and considered helpful for MSM.^51^ Similarly, young people visiting youth clinics expressed the importance of HCPs’ confidentiality and that they are used to and only work with young people.^5^ Trust in HCPs’ confidentiality was also described as important in the process of men disclosing victimization.^28^

#### 4. Healthcare system factors

The healthcare system influenced the HCP-user relationship through its effects on HCPs and the healthcare encounters. Among other issues, the literature discussed the organization of healthcare, the holistic approach (or the lack of it), SRHC as traditionally women-centred care and men’s SRHC as “nobody’s mandate”.

##### 4.1. Men’s SRH is not a priority

The literature indicated that the clinical training and organization of care does not give men’s SRH enough priority. Nurses, for example, highlighted the lack of basic medical training and organizational support to deal with men’s sexual health issues. Their main source of knowledge about men’s SRH was received from pharmaceutical companies.^16^ Similarly, midwifery education and clinical training doesn’t regularly include andrology, which together with lack of time and organizational support hindered them from providing counselling to men.^17^

Another example of the low priority of men’s SRH was the lack of follow-up and continuity of care, which was reported in various services. For example, men reported not being followed-up after being prescribed medication for sexual function.^27^ Additionally, the stays of men with the family after delivery was not welcomed in some hospitals, even though this was important for men in order to feel supported and to support their new family.^46^

The lack of prioritizing men’s SRH was also reflected by the few prevention activities that healthcare performs regarding men’s SRH. For example, the vast majority of MSM did not encounter any HIV/STI prevention services, despite the importance of making it more available.^70^ Another example was the missed opportunity to counsel for sexual health in around one-third of men testing for HIV.^50^

##### 4.2. Lack of holistic care

The literature discussed the lack of holistic care in SRH services. For example, psychological aspects of infertility were usually not acknowledged and therefore overlooked.^35^ For couples with repeated pregnancy loss, psychological counselling was restricted to a few with certain criterion and also without considering individual situations.^42^ Furthermore, antenatal care was perceived to focus mainly on medical support and rarely on emotional and psychological support, leaving only few users being very satisfied with this aspect of antenatal care.^14,74^ Consequently, men who were subjected to gender-based violence were less likely to seek help unless they had severe physical injuries.^28^

##### 4.3. Women-centred reproductive healthcare, a compromised right for inclusion of men

Both men and women expressed a wish to include men and to focus on “the couple” rather than one partner, that is, equal partners sharing a common reason for visiting reproductive healthcare.^15,35,42,52^ Even though men felt that the focus on women in reproductive healthcare is reasonable, they stated that this attention should not exclude men.^35,48^ The feeling of exclusion was experienced by men in different reproductive health services, including fertility and antenatal care.^14,15,37,48^ One study showed that the investigations and treatments focused only on the women, even when the cause of infertility was a low sperm count, which led to perceive infertility care as the “women’s world”.^15^ Additionally, the midwives discussed sexual and reproductive rights for men as being women’s partners rather than being men’s own rights, and men’s concerns about contraception are dependent upon his partner’s choice to include him or not in contraceptive counselling.^17^

##### 4.4. Men’s SRH is nobody’s responsibility

Different HCPs expressed their concern about men’s sexual health as “no one’s responsibility”. The attitudes of midwives toward providing counselling to men were divided. Some were positive and found it a continuation of their current responsibilities that concerned women.^17^ This opinion was shared by men of pregnant partners who expressed their trust and faith in midwives and saw them as the best ones to promote sexual health among men.^48^ Other midwives were reluctant and expressed their difficulty in providing counselling to men. For them, the pregnancy is about the woman’s body, and thus, man’s participation was not evident.^17^ Nurses also questioned if men’s sexual health is their duty, especially if it included an emotional aspect. In their opinion, primary care was not equipped to deal with sexual health problems; therefore, they often referred patients to other healthcare units.^16^

#### 5. Sociopolitical factors

The outer layer of the model (Figure 4) discusses the social and political factors, including social and gender norms, which affect the healthcare system and the attitudes and behaviours of HCPs and users.

##### 5.1. Social and gender norms

The literature described traditional social and gender norms as contributing to setting values for men that hinder their abilities to cope or seek help and affect their sexual health and wellbeing. For example, infertility was described as a “malfunction of manhood” that is faced by denial and changes how men perceive their masculinities. The absence of sperm was an identity question and a threat to men’s masculinities.^15,26,31,67^ Similarly, suffering the decline in sexual function associated with a diagnosis of prostate cancer was perceived as threatening to the male identity and therefore accompanied by feelings of inadequacy and not being a “real man”.^27,77^

Furthermore, the attempts of men to conform to traditional masculinity norms affected their ability to talk about experiences of violence, especially if they were exposed to intimate partner violence.^28^ Additionally, transmen had experiences of vulnerability during gynaecological examination or when they resumed menstrual bleeding after family planning treatment. This was perceived as stressful, humiliating and uncomfortable, as well as a reminder of a sex they “wanted to forget”.^83^

These “threatened masculinities” were also reflected through men’s health seeking behaviours. Men disregarded their sexual health, delayed admission of the problem and opted to distance themselves from seeking healthcare.^13^ For example, young men had more difficulties to admit their SRH needs and to seek help as compared to young women.^5^ Similarly, the midwives indicated that men only seek help when they have severe symptoms, while also noting that young men are increasingly attending STI testing and are more open to discuss sexual health.^17^

Men expressed increased social expectations on them to be more involved in healthcare during pregnancy and birth, which corresponded to personal willingness and desire to share responsibility for the security and support of their partners.^27,38,44,46^ Men were also eager to participate in other reproductive healthcare services, such as infertility treatment and home abortion.^35,71,82^ However, men were faced with barriers in their desire to participate and experienced “paddling upstream” to fulfil their involvement.^39^

The literature also discussed how social and gender norms affect the healthcare system being perceived as women-centred. Youth clinics, for example, were perceived as a place for the SRH of girls, which created a barrier for young men seeking healthcare.^5^ Additionally, the social norms hindered HCPs talking about sexual health, especially when the patient is older than the HCP.^16^ In turn, HCPs reinforced these social norms by supporting the traditional gender expectations of the woman as the primary infant caregiver and overlooking the importance of shared parenthood and including the man in infant care.^38,84^

Studies also described how social and gender norms affected the way healthcare deals with victimized men. The training and education the emergency departments offered in Sweden about caring for violence victims focuses only on women and children and not victimized men.^85^ Similar experiences of the reinforcement of traditional gender positions by HCPs were perceived by men subjected to intimate partner violence. These men felt alone since society did not acknowledge their experiences, and the HCPs expected them to embody traditional ideals of masculinities.^28^

##### 5.2. Policies

Politics and policies were rarely discussed in the literature, but there were some mentions of the regulations and guidelines in SRHC, which have been discussed under point 4. The only mention of policy was in the context of gender-based violence. While most of the counties and emergency departments in Sweden had a policy about the care for victims of violence, these policies focus merely on women and children but not men or other groups.^85^

## Discussion

In the previous section we reviewed, charted and synthesized the available literature in relation to the factors influencing men’s experiences in SRHC. To summarize, the majority of the reviewed literature discussed men’s experiences in reproductive healthcare, mainly care related to infertility, pregnancy and birth. The literature lacked men’s perspectives on contraception, including condom use and vasectomy. Regarding sexual healthcare, the available literature captured mainly STIs and HIV treatment and prevention but not men’s experiences in other sexual health issues, such as impotence or gender-based violence. The literature also lacked the perspectives of particular groups of men, such as Indigenous, national minorities and men with functional variations. Furthermore, migrants and MSM were almost only mentioned in relation to HIV treatment and prevention.

The literature indicated that men face difficulties to be included in reproductive healthcare, where they are mostly treated as an accompanying partner, receiving little attention. The knowledge and attitudes of HCPs were crucial for their ability to discuss men’s SRH and also for men’s experiences in SRHC. Furthermore, the literature rarely discussed healthcare organization and policies and how they affect men’s health seeking behaviours and experiences in SRHC. Lastly, men’s right to SRH is usually not stressed in the literature, unless it is related to a specific group of men, such as MSM and transmen.

While we presented the factors influencing men’s experiences in SRHC in separate levels and the reviewed articles did not explicitly study the interaction between these levels, the theoretical framework still enables us to understand the interaction between these determinants. We presented some examples in the results of how these levels are linked and influence each other. The interaction between gender and social norms with the other determinants might be of special significance. For example, the literature described how traditional social and gender norms affect the attitudes and behaviours of HCPs. A clear example of this was how men were treated as an accompanying partner during healthcare visits related to infertility, antenatal care and birth. In many cases men are still not seen as an equal partner or as a primary caregiver for their new-borns, which likely influences the attitudes of HCPs toward men seeking antenatal care, in turn affecting men’s experiences of those services negatively.^14,39^

While traditional gender norms and values of masculinities provide important pieces in explaining men’s health seeking behaviours, a more comprehensive picture of men’s experiences in SRHC is needed. The literature showed other determinants of men’s experiences in SRHC, including how the healthcare system is organized. It seems SRHC in the Nordic countries focuses mainly on women, while there is a lack of knowledge about men’s SRH and no clear entry for men into SRHC. The healthcare system should adapt a gender-responsive approach that ensures accessible healthcare services for men and which through its approach addresses the impacts of gender norms on men, women and HCPs.^2,86^

To reach universal access to SRHC and gender equity, it is of importance to engage men in SRH and ensure that their needs are met.^87^ Improving men’s experiences in SRHC in the Nordic countries is not only important for improving men’s SRH but also could enable men to strengthen their support of women’s SRH and thus gender equality.^87,88^ Meeting men’s needs for SRHC could consequently decrease STIs, unintended pregnancies and improve parenting and family relationships.^9,21^ Such specific focus on men in the SRHC organization to improve men’s health and rights with the goal to contribute to gender equality will benefit both men and women.^89,90^

### Strengths and limitations of this study

To the best of our knowledge, this review is the first to examine the experiences of men in SRHC in the Nordic countries. The review provides interesting and important information about these experiences, by organizing them in a theoretical framework that make it easier to understand and draw conclusions.

Even though we developed and followed our search strategies thoroughly, the review has some limitations. The broad nature of the field and the wide variety of terms related to SRH make it difficult to assure the inclusion of all relevant literature. Additionally, due to practical reasons, the search was restricted to two databases but complemented with manually screening the reference lists of the identified literature. Another strength of this review was the use of a Nordic-specific database without restriction to language, which ensured an equal inclusion of the literature from other Nordic countries, even though most of the literature in this review was published in Sweden.

Furthermore, the adapted framework allowed us to use a relevant ecological lens on men’s experiences in SRHC and to systematically identify and categorize the concepts discussed in the selected literature. However, the use of this framework might have caused us to overlook aspects of the research topic that fell outside the interest of this scoping review.

Finally, it is important to note that, when reporting results and discussing them, we choose to implicitly treat the Nordic countries as essentially similar. While we argue that this makes sense because of the actual similarities between these countries and their healthcare systems, we also acknowledge that this might obscure important differences between or within countries (e.g., relation to place of residence (rural vs. urban) or cultural differences).

## Conclusion

Despite the uncontroversial importance of men’s right to access SRHC on equal terms, the available literature indicated that SRH is mainly the domain of women and healthcare around men’s SRH is not sufficiently prioritized. A more comprehensive picture of men’s experiences in SRHC is needed.

There is a lack of knowledge about men’s SRH and no clear entry for men into SRHC. This indicates the necessity for improvements in the medical education of HCPs and in health system interventions. Further research should examine the influence of policies and the healthcare organization on men’s access and experiences in SRHC and explore the identified knowledge gaps of men’s experiences in SRHC related to sexual function, contraceptive use and gender-based violence.

## Data Availability

The research has no primary collected data

## Author contributions

MB was the lead reviewer and author of the manuscript. AKH and JPS contributed to the identification and selection of articles. All authors contributed to data interpretation. All authors provided comments and agreed on the final version of the manuscript. AKH was the project leader.

## Appendices

**Appendix 1:**
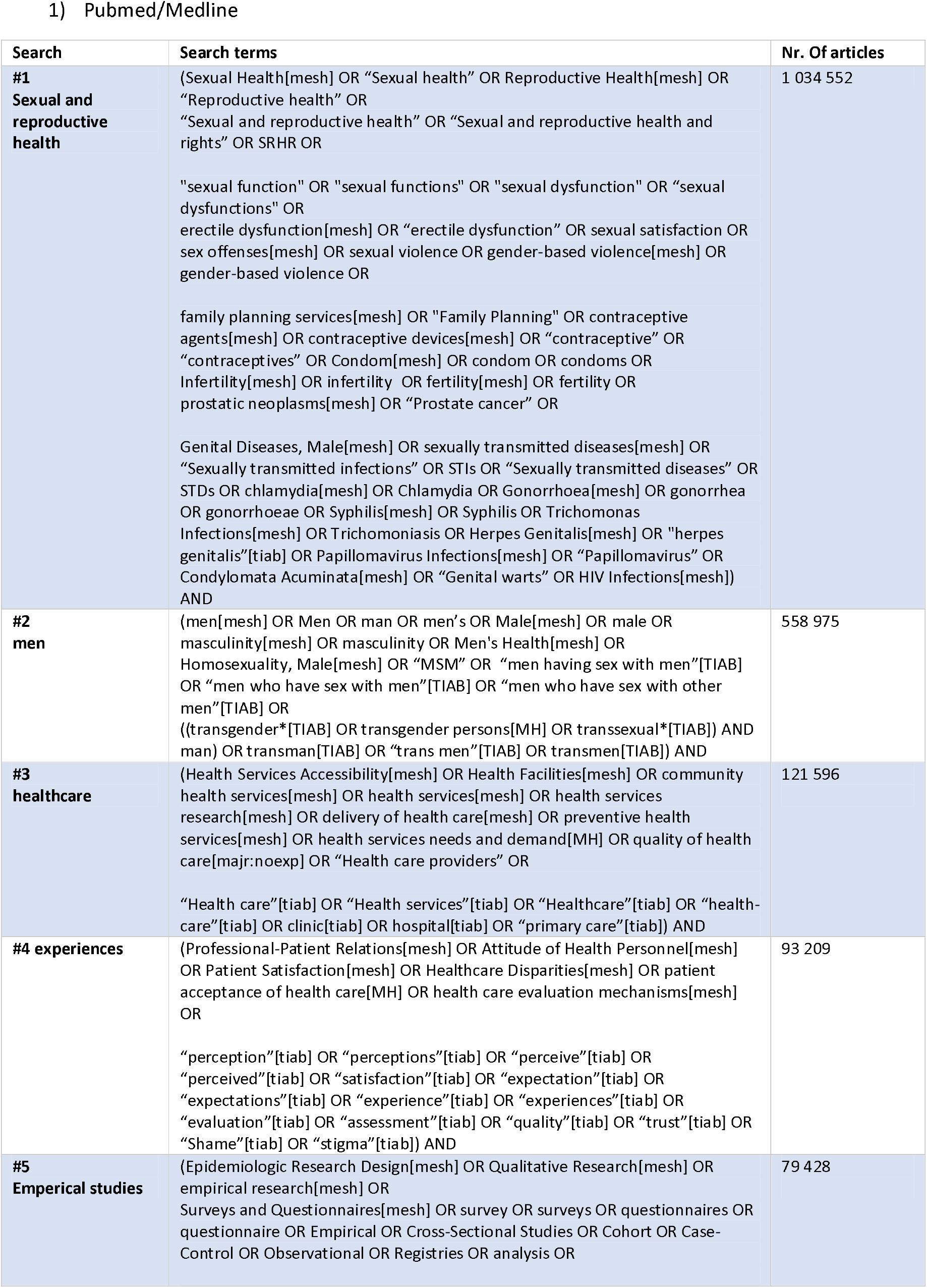
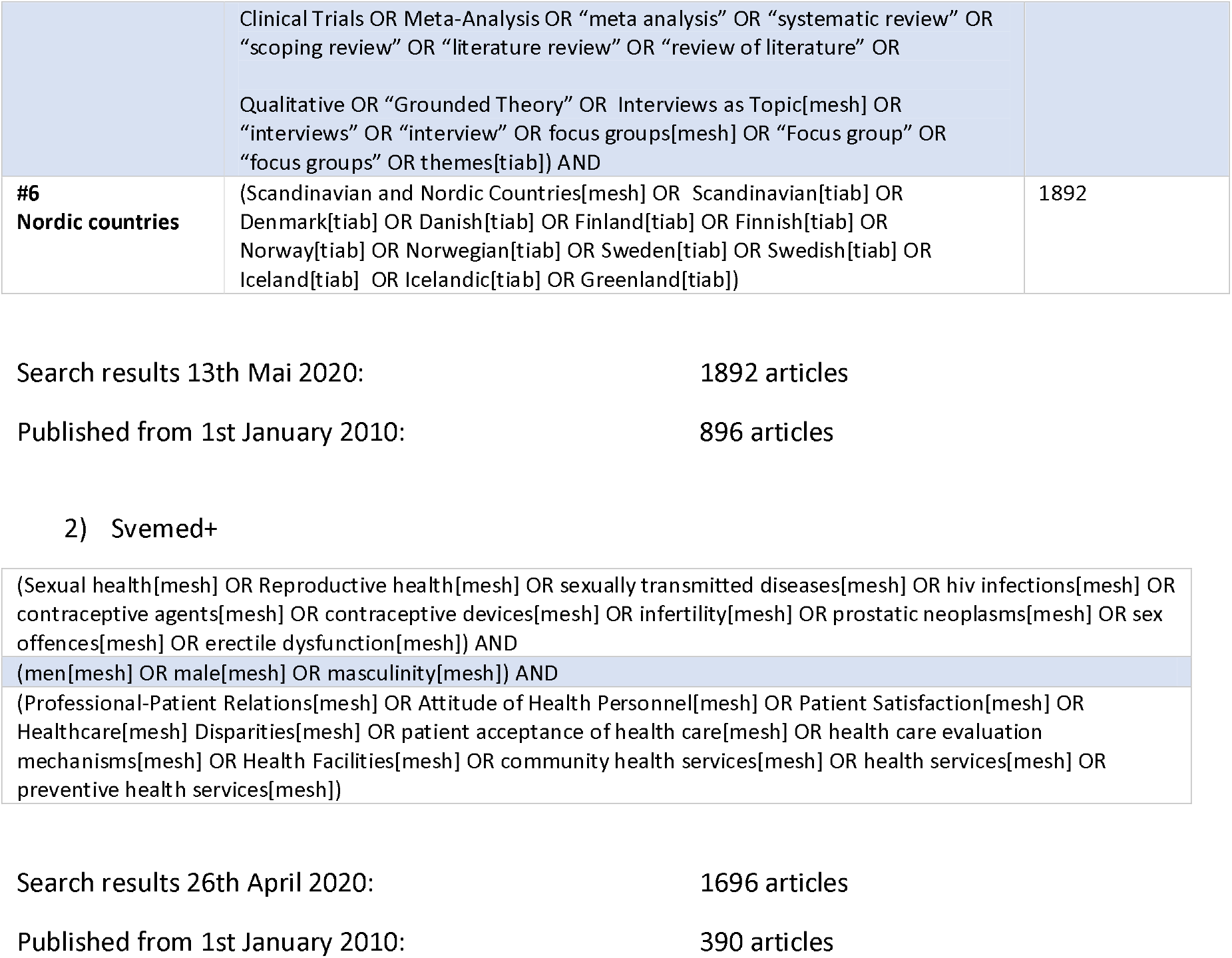
Söktermer.

1) Pubmed/Medline
| Search | Search terms | Nr. Of articles |
| --- | --- | --- |
| **#1**<br>**Sexual and reproductive health** | (Sexual Health[mesh] OR "Sexual health" OR Reproductive Health[mesh] OR "Reproductive health" OR "Sexual and reproductive health" OR "Sexual and reproductive health and rights" OR SRHR OR<br>"sexual function" OR "sexual functions" OR "sexual dysfunction" OR "sexual dysfunctions" OR<br>erectile dysfunction[mesh] OR "erectile dysfunction" OR sexual satisfaction OR sex offenses[mesh] OR sexual violence OR gender-based violence[mesh] OR gender-based violence OR<br>family planning services[mesh] OR "Family Planning" OR contraceptive agents[mesh] OR contraceptive devices[mesh] OR "contraceptive" OR "contraceptives" OR Condom[mesh] OR condom OR condoms OR Infertility[mesh] OR infertility OR fertility[mesh] OR fertility OR prostatic neoplasms[mesh] OR "Prostate cancer" OR<br>Genital Diseases, Male[mesh] OR sexually transmitted diseases[mesh] OR "Sexually transmitted infections" OR STIs OR "Sexually transmitted diseases" OR STDs OR chlamydia[mesh] OR Chlamydia OR Gonorrhoea[mesh] OR gonorrhea OR gonorrhoeae OR Syphilis[mesh] OR Syphilis OR Trichomonas Infections[mesh] OR Trichomoniasis OR Herpes Genitalis[mesh] OR "herpes genitalis"[tiab] OR Papillomavirus Infections[mesh] OR "Papillomavirus" OR Condylomata Acuminata[mesh] OR "Genital warts" OR HIV Infections[mesh]) AND | 1 034 552 |
| **#2**<br>**men** | (men[mesh] OR Men OR man OR men's OR Male[mesh] OR male OR masculinity[mesh] OR masculinity OR Men's Health[mesh] OR Homosexuality, Male[mesh] OR "MSM" OR "men having sex with men"[TIAB] OR "men who have sex with men"[TIAB] OR "men who have sex with other men"[TIAB] OR<br>((transgender*[TIAB] OR transgender persons[MH] OR transsexual*[TIAB]) AND man) OR transman[TIAB] OR "trans men"[TIAB] OR transmen[TIAB]) AND | 558 975 |
| **#3**<br>**healthcare** | (Health Services Accessibility[mesh] OR Health Facilities[mesh] OR community health services[mesh] OR health services[mesh] OR health services research[mesh] OR delivery of health care[mesh] OR preventive health services[mesh] OR health services needs and demand[MH] OR quality of health care[majr:noexp] OR "Health care providers" OR<br>"Health care"[tiab] OR "Health services"[tiab] OR "Healthcare"[tiab] OR "health-care"[tiab] OR clinic[tiab] OR hospital[tiab] OR "primary care"[tiab]) AND | 121 596 |
| **#4 experiences** | (Professional-Patient Relations[mesh] OR Attitude of Health Personnel[mesh] OR Patient Satisfaction[mesh] OR Healthcare Disparities[mesh] OR patient acceptance of health care[MH] OR health care evaluation mechanisms[mesh] OR<br>"perception"[tiab] OR "perceptions"[tiab] OR "perceive"[tiab] OR "perceived"[tiab] OR "satisfaction"[tiab] OR "expectation"[tiab] OR "expectations"[tiab] OR "experience"[tiab] OR "experiences"[tiab] OR "evaluation"[tiab] OR "assessment"[tiab] OR "quality"[tiab] OR "trust"[tiab] OR "Shame"[tiab] OR "stigma"[tiab]) AND | 93 209 |
| **#5**<br>**Emperical studies** | (Epidemiologic Research Design[mesh] OR Qualitative Research[mesh] OR empirical research[mesh] OR<br>Surveys and Questionnaires[mesh] OR survey OR surveys OR questionnaires OR questionnaire OR Empirical OR Cross-Sectional Studies OR Cohort OR Case-Control OR Observational OR Registries OR analysis OR | 79 428 |
|  | Clinical Trials OR Meta-Analysis OR "meta analysis" OR "systematic review" OR "scoping review" OR "literature review" OR "review of literature" OR<br>Qualitative OR "Grounded Theory" OR Interviews as Topic[mesh] OR "interviews" OR "interview" OR focus groups[mesh] OR "Focus group" OR "focus groups" OR themes[tiab] AND |  |
| **#6**<br>**Nordic countries** | (Scandinavian and Nordic Countries[mesh] OR Scandinavian[tiab] OR Denmark[tiab] OR Danish[tiab] OR Finland[tiab] OR Finnish[tiab] OR Norway[tiab] OR Norwegian[tiab] OR Sweden[tiab] OR Swedish[tiab] OR Iceland[tiab] OR Icelandic[tiab] OR Greenland[tiab]) | 1892 |

2) Svemed+
|  |
| --- |
| (Sexual health[mesh] OR Reproductive health[mesh] OR sexually transmitted diseases[mesh] OR hiv infections[mesh] OR contraceptive agents[mesh] OR contraceptive devices[mesh] OR infertility[mesh] OR prostatic neoplasms[mesh] OR sex offences[mesh] OR erectile dysfunction[mesh]) AND |
| (men[mesh] OR male[mesh] OR masculinity[mesh]) AND |
| (Professional-Patient Relations[mesh] OR Attitude of Health Personnel[mesh] OR Patient Satisfaction[mesh] OR Healthcare[mesh] Disparities[mesh] OR patient acceptance of health care[mesh] OR health care evaluation mechanisms[mesh] OR Health Facilities[mesh] OR community health services[mesh] OR health services[mesh] OR preventive health services[mesh]) |

Search results 26th April 2020: 1696 articles
Published from 1st January 2010: 390 articles

**Appendix 2:**
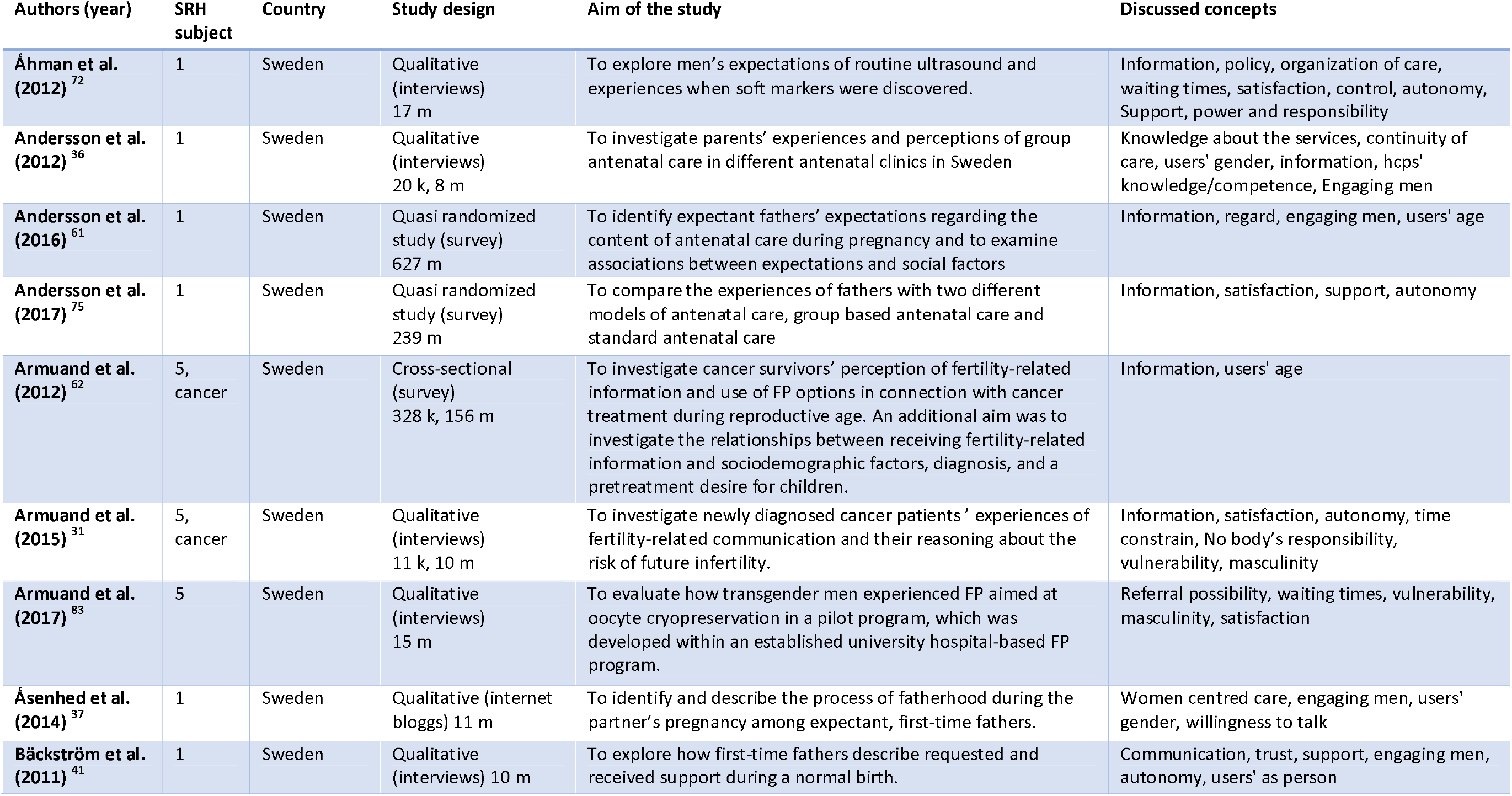
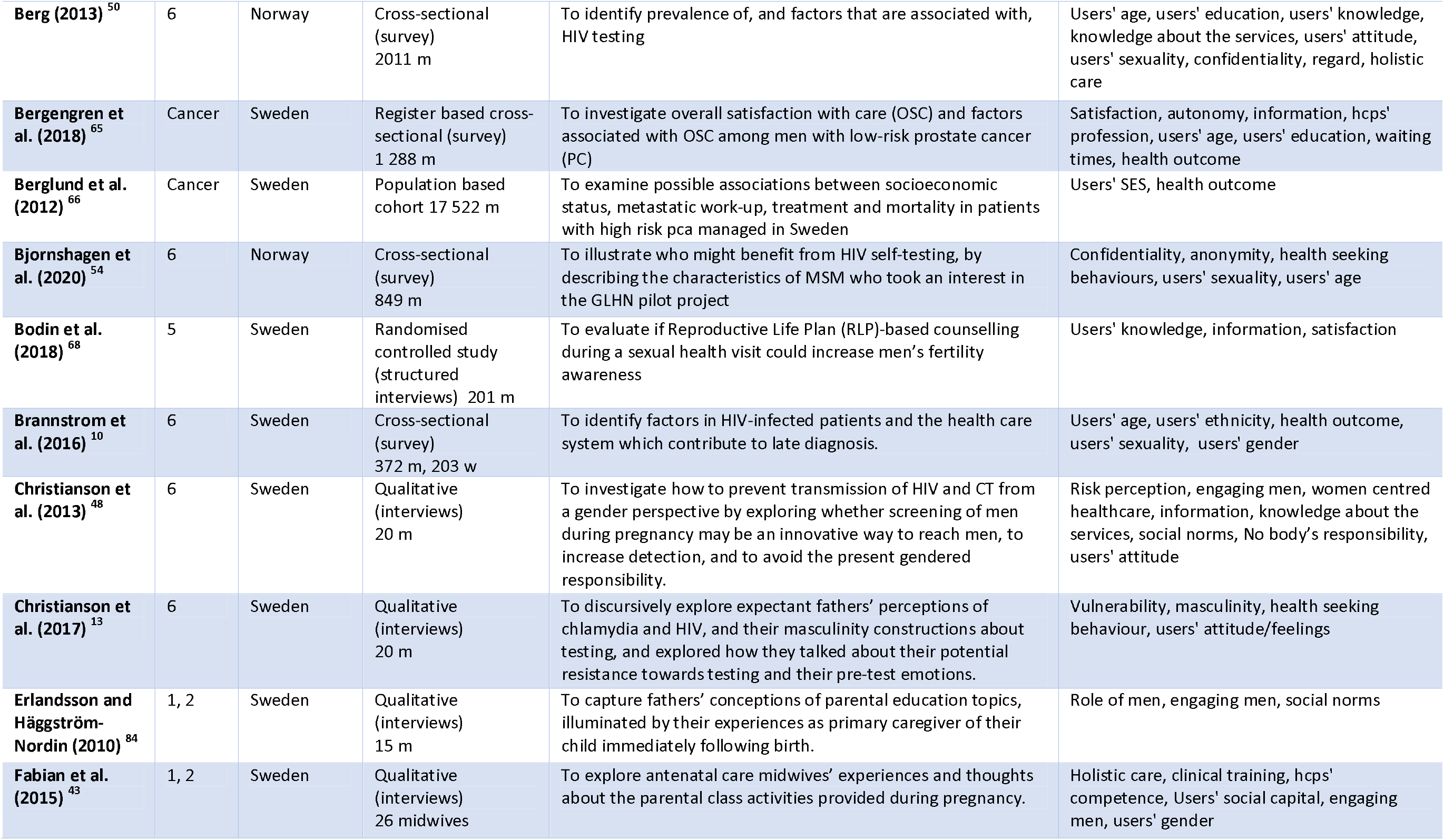
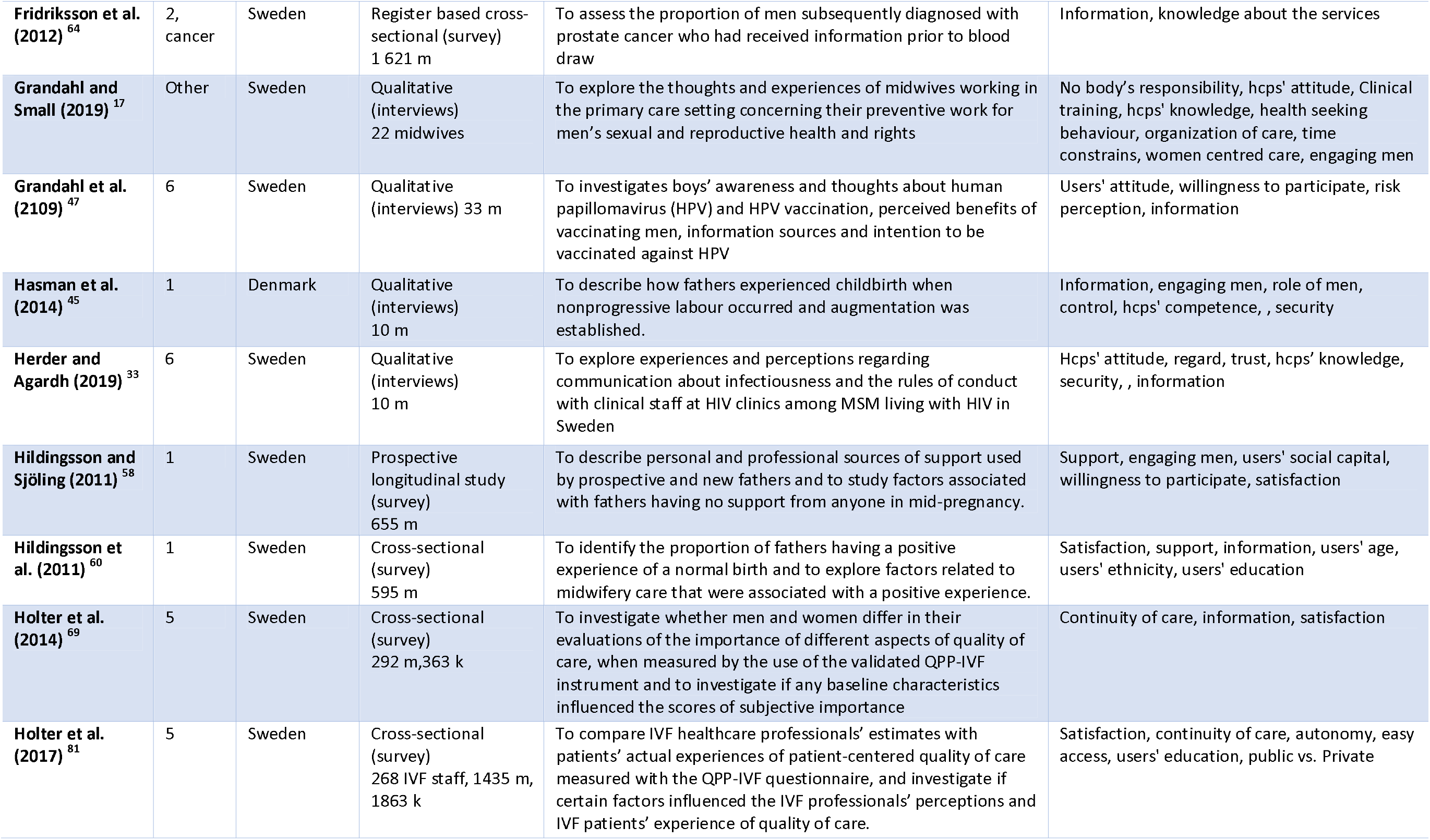
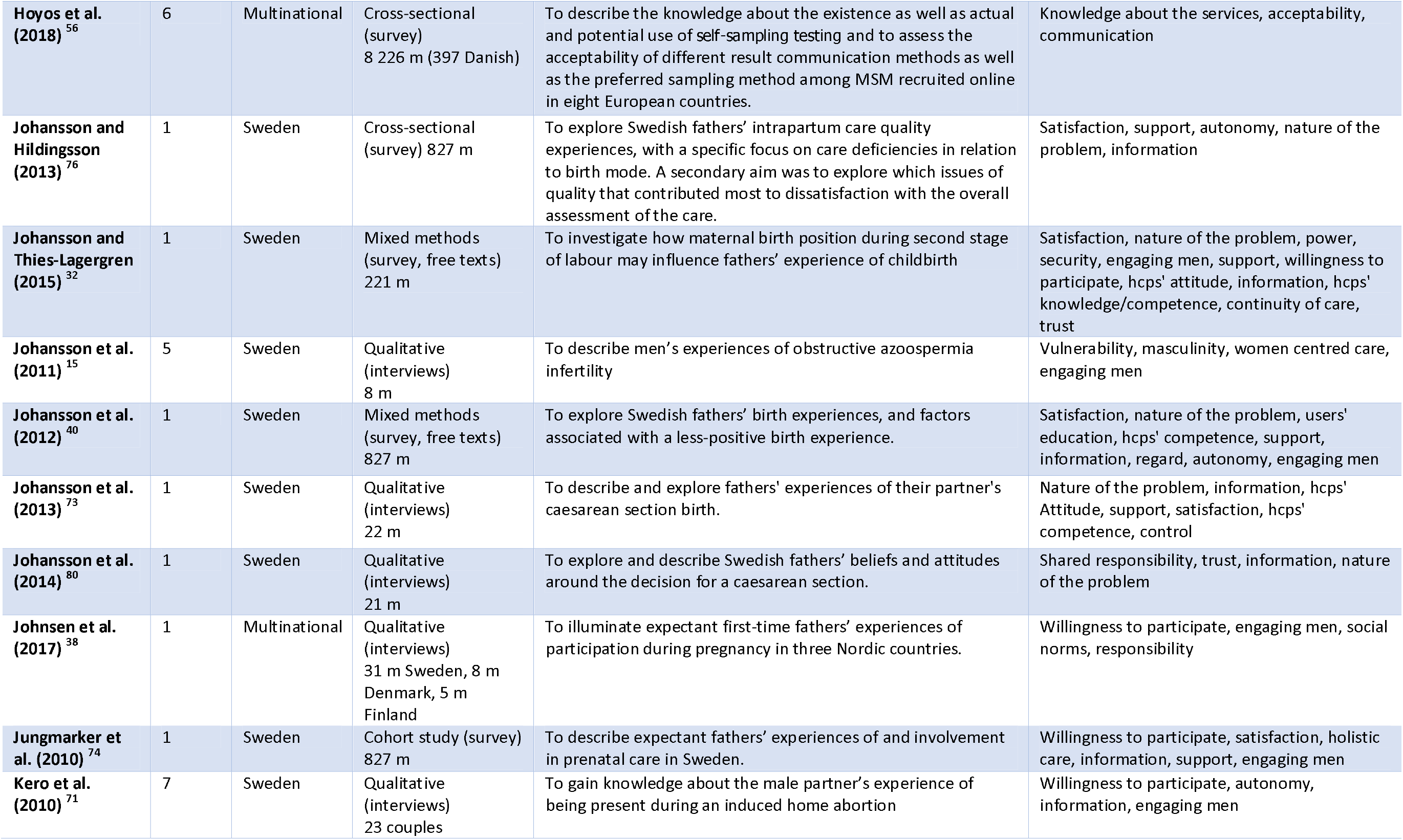
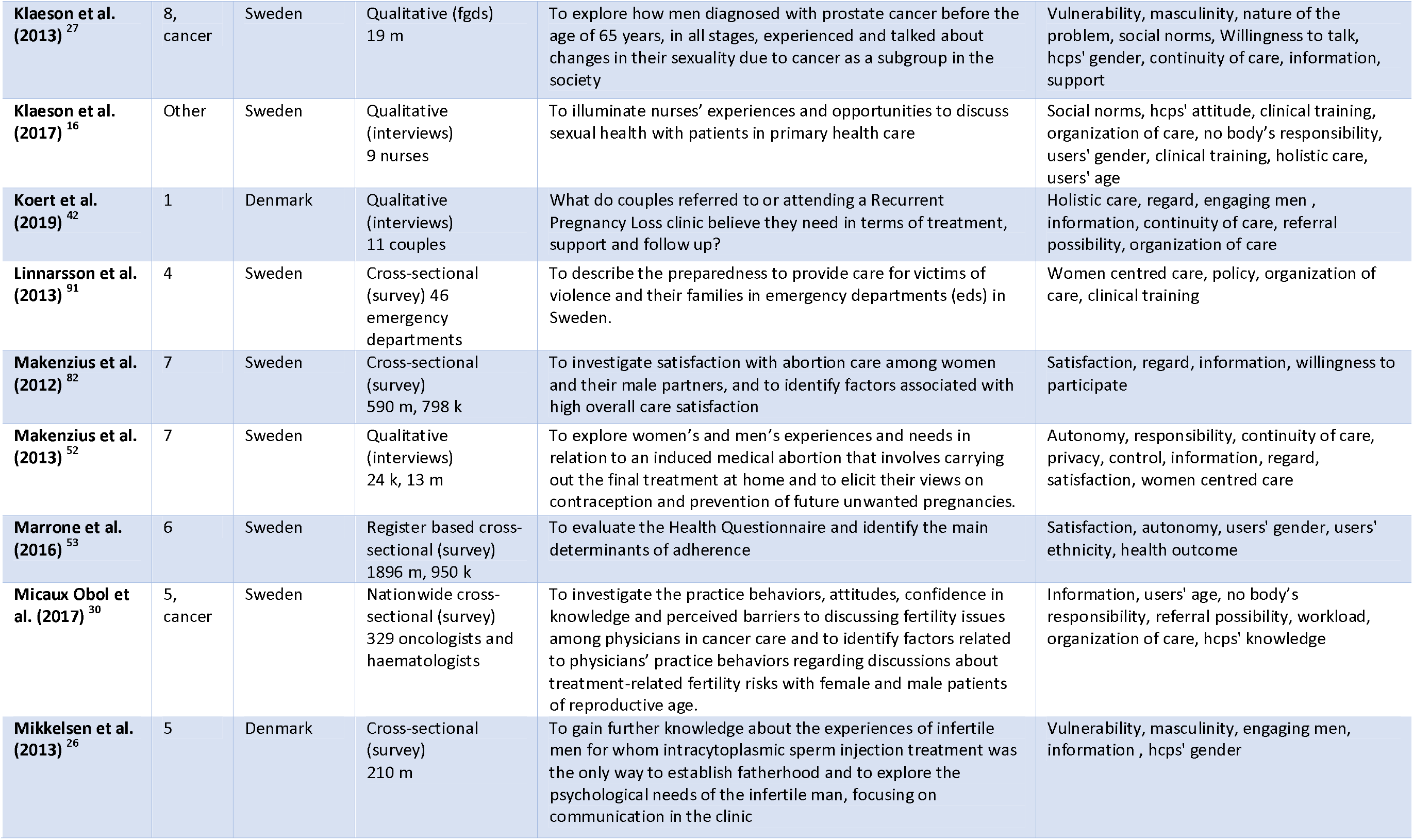
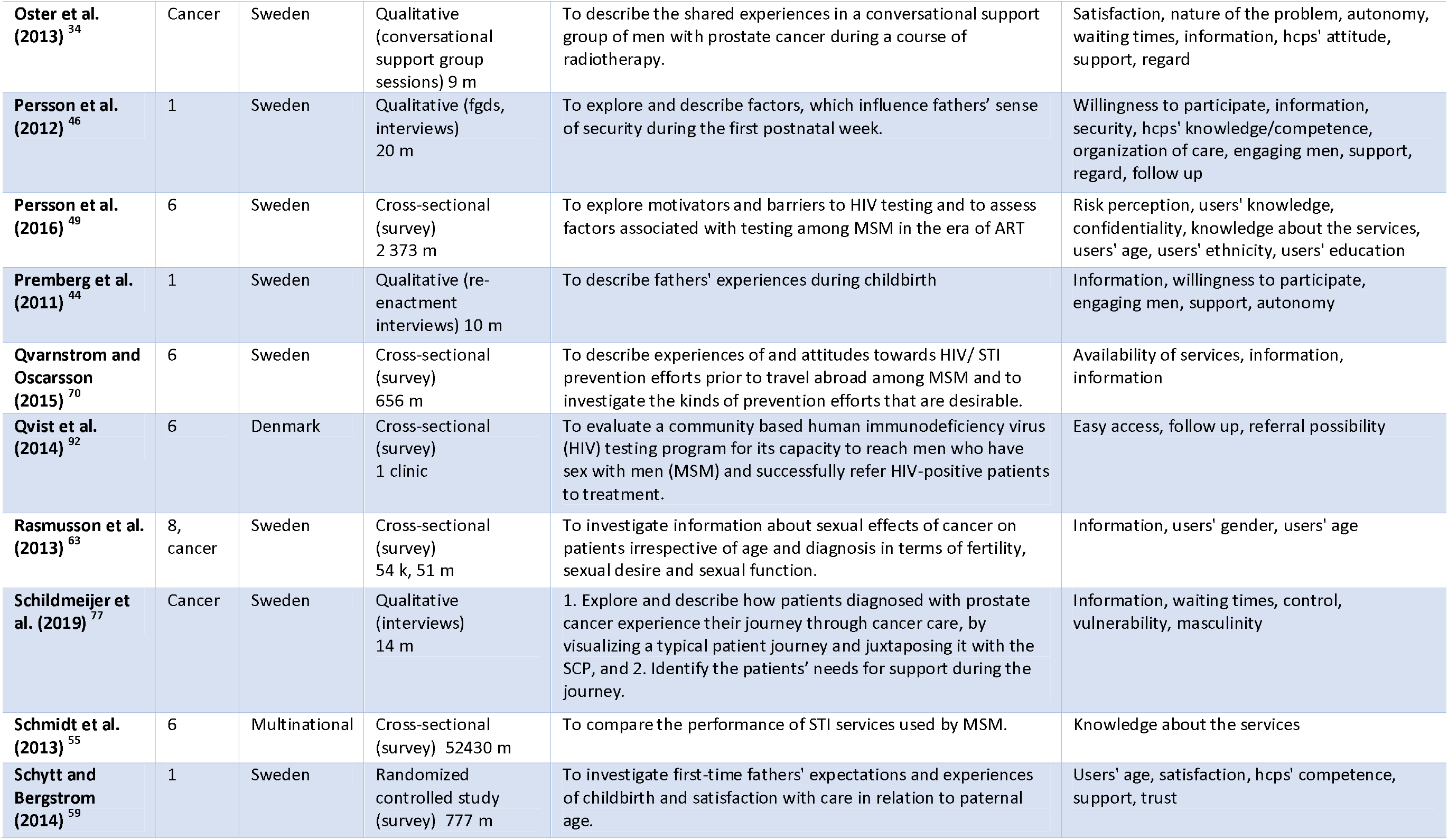
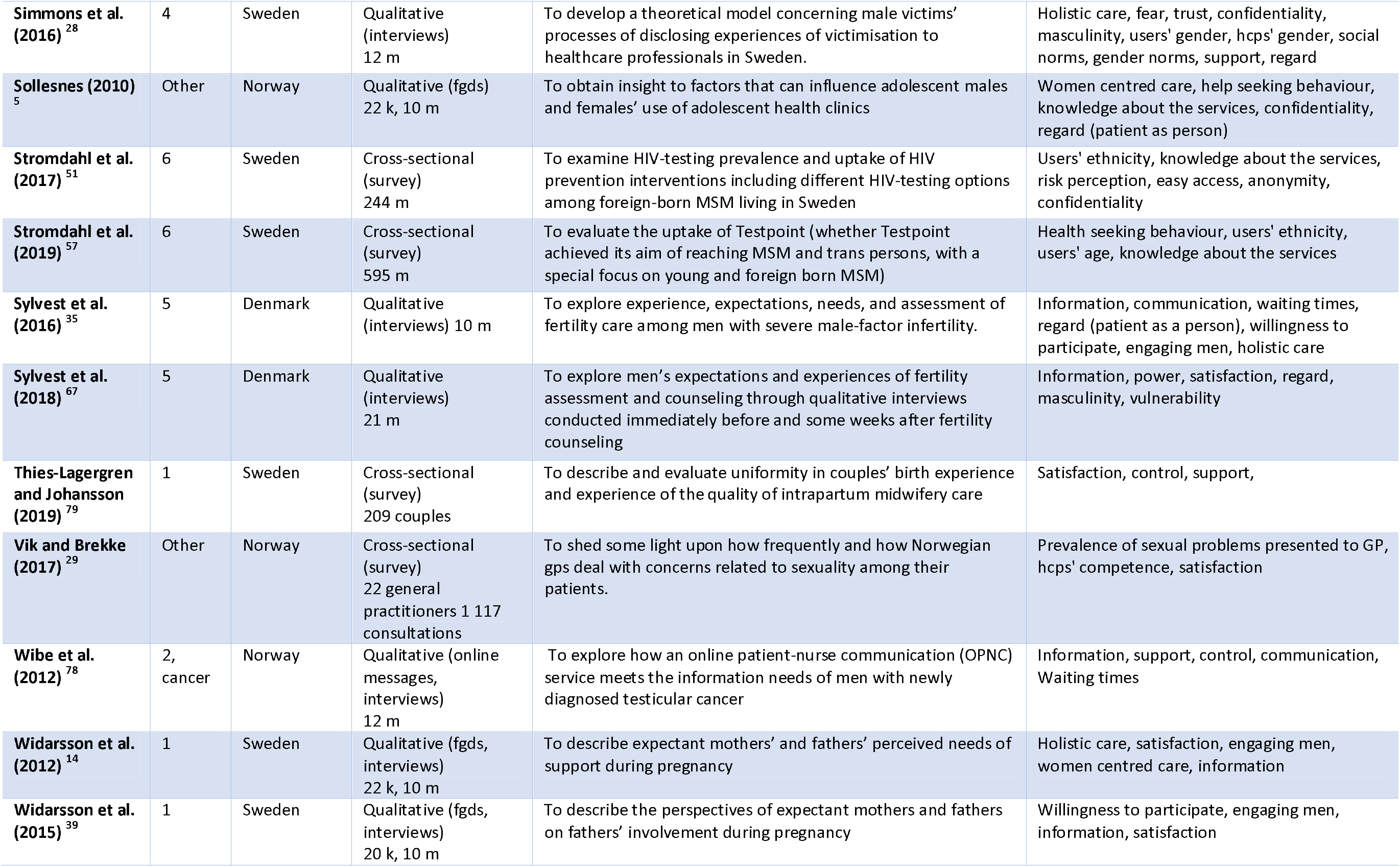
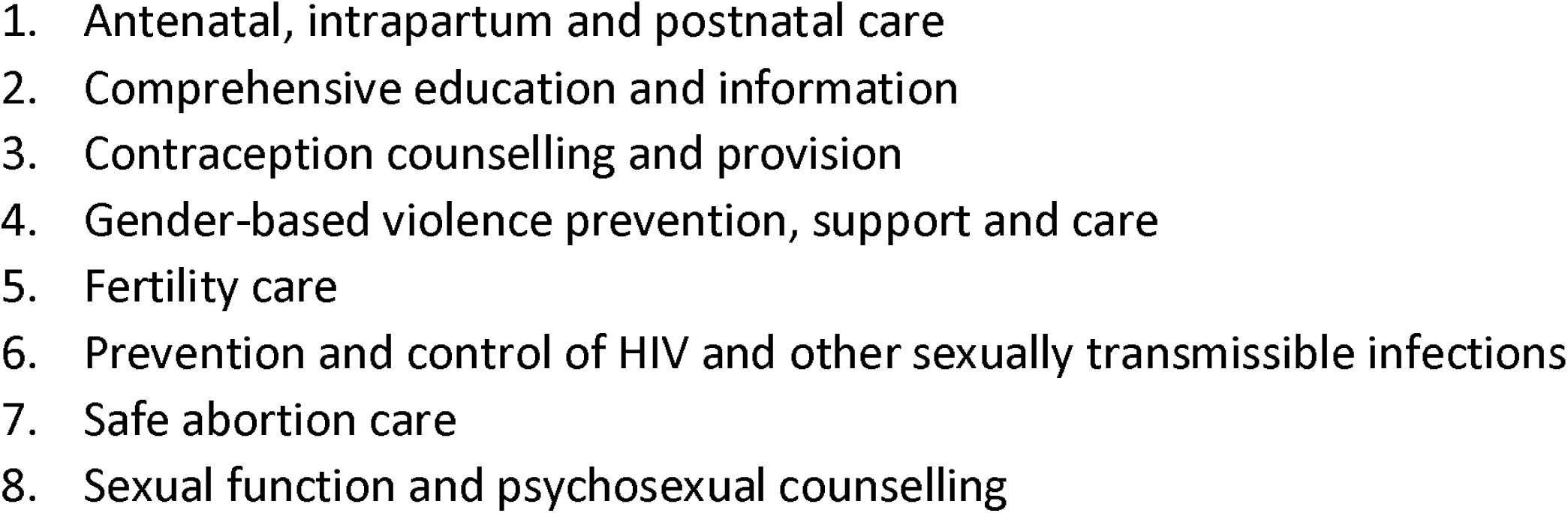
lista över inkluderade artiklar om mäns erfarenheter inom sexuell och reproduktiva hälso och sjukvård i de nordiska länderna.

